# Sensitivity of commercial Anti-SARS-CoV-2 serological assays in a high-prevalence setting

**DOI:** 10.1101/2020.06.11.20128686

**Authors:** Lisa Müller, Philipp N. Ostermann, Andreas Walker, Tobias Wienemann, Alexander Mertens, Ortwin Adams, Marcel Andree, Sandra Hauka, Nadine Lübke, Verena Keitel, Ingo Drexler, Veronica Di Cristanziano, Derik Franz Hermsen, Rolf Kaiser, Friedrich Boege, Florian Klein, Heiner Schaal, Jörg Timm, Tina Senff

**Affiliations:** Institute of Virology, University Hospital Düsseldorf, Heinrich Heine University Düsseldorf, Düsseldorf, Germany; Institute of Medical Microbiology and Hospital Hygiene, Heinrich Heine University Düsseldorf, Düsseldorf, Germany; Department of Gastroenterology, Hepatology and Infectious Diseases, University Hospital Düsseldorf, Heinrich Heine University Düsseldorf, Düsseldorf, Germany; Institute of Virology, University of Cologne, Faculty of Medicine and University Hospital of Cologne, Cologne, Germany; Institute of Clinical Chemistry and Laboratory Diagnostics, Medical Faculty, University Düsseldorf, Düsseldorf, Germany

**Keywords:** SARS-CoV-2, COVID-19, serology, neutralising antibodies, immunofluorescence test

## Abstract

We analysed SARS-CoV-2 specific antibody responses in 42 social and working contacts of a super-spreader from the Heinsberg area in Germany. Consistent with a high-prevalence setting 26 individuals had SARS-CoV-2 antibodies determined by in-house neutralisation testing. These results were compared with four commercial assays, suggesting limited sensitivity of the assays in such a high-prevalence setting. Although SARS-CoV-2 nucleocapsid-restricted tests showed a better sensitivity, spike-based assays had a stronger correlation with neutralisation capacity.

---

Sensitive serological high throughput SARS-CoV-2 assays are of great importance for seroprevalence studies and retrospective diagnosis of SARS-CoV-2 infections. They could aid in identifying donors for convalescent plasma therapy and determine antibody titres to assess possible immunity after vaccination [1].

Here, we assessed the performance characteristics of several serological tests for SARS-CoV-2 antibodies, including a neutralisation test within a high-risk cohort.

## Social and working network contacts of the index patient

On 24^th^ February 2020, a patient from the Heinsberg district with no known travel history to SARS-CoV-2 risk areas was diagnosed by PCR with SARS-CoV-2. Contact tracing identified 37 secondary cases by February 28^th^, and >1,000 linked to a super-spreading event held on 15^th^ February 2020 [2]. Here, we analysed blood samples of 42 social and working network contacts of the index patient collected on April 9, 2020. The study population contains slightly more females than males (26/16 61.9%, 38.1%) and individuals were aged between 18 and 70 years (median 44). Despite only eight of the 42 individuals were tested positive for SARS-CoV-2-RNA, 26 described symptoms including fever (38.5%), cough (65.4%), fatigue (50%), shortness of breath or difficulty of breathing (30.8%) while 16 reported no symptoms (table S1). To assess the performance of several serological tests and to identify past infections, we analysed SARS-CoV-2 specific antibody responses in sera from the 42 individuals.

## SARS-CoV-2 neutralising antibody levels

Neutralising antibodies with neutralisation test (NT) titres ≥20 were detected in 26 of 42 serum samples (61.9%). Besides the RT-PCR confirmed SARS-CoV-2 cases (n=8), 13 out of 19 symptomatic (68.4%) and 5 of 15 asymptomatic (33.3%) individuals had neutralising antibodies (table S2). Neutralising antibody levels in asymptomatic individuals were significantly lower compared to PCR confirmed cases (p≤0.01, figure 1). To assess the presence of Anti-SARS-CoV-2 IgG, an immunofluorescence test (IFT) was performed (supplementary text). Of the 26 sera positive in the neutralisation assay, 23 were also positive in the IFT (sensitivity 88.5% CI95% [0.710-0.960]) and negative IFT results were associated with low (≤40) NT-titres.

**Figure 1:**
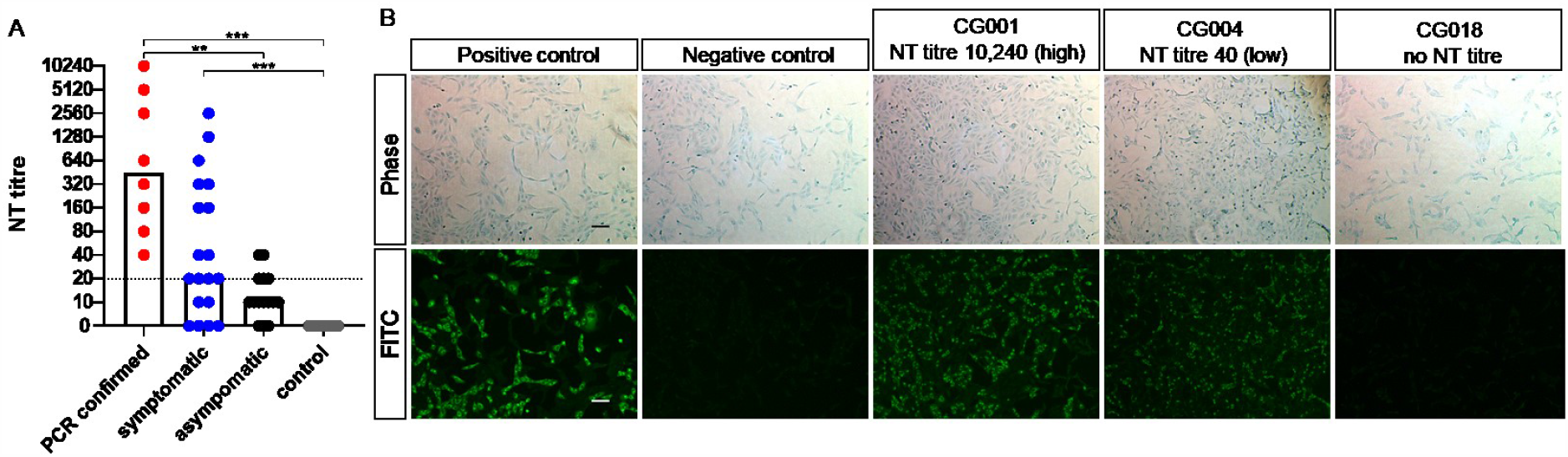
SARS-CoV-2 neutralising antibodies stratified according to status of study participants and exemplary immunofluorescence test results, Germany Heinsberg District, April 2020 (n=42) FITC: Fluorescein isothiocyanate; NT: neutralisation test; (A) Neutralisation assay results of 42 individuals grouped by their status in PCR positive (red), symptomatic (blue) and asymptomatic (black) and 11 control sera from healthy individuals sampled before December 2019. The reciprocal of the NT titre is depicted, and bars represent the respective median. The cut off was defined as ≥20. One-way ANOVA was used to compare groups **p≤0.01 ***p≤0.001 (B) Exemplary Anti-SARS-CoV-2 IgG immunofluorescence test results of 3 out of 42 tested individuals. Phase contrast (a-e) and FITC fluorescence detected at 488nm (f-j). Serum of a severe hospitalised COVID-19 case served as a positive control (a+f) and (b+g) depict the result of a negative control serum. IFT results from a patient with a high NT titre (c+h; NT titre10,240), a low NT titre (d+i NT titre 40) and no neutralisation potential (e+j). Scale bar is 100 µm.

## Sensitivity of commercial high-throughput SARS-CoV-2 antibody assays

Based on IFT and NT results 26 sera were defined as SARS-CoV-2 antibody positive. Since both methods are time consuming and labour-intensive, the suitability of antibody testing was analysed with four different commercially available automated serological test systems targeting either the nucleocapsid protein (N) or the spike protein (S) of SARS-CoV-2 (supplementary text) (Table 1, Figure S1/S2). Our study included the (i.) EUROIMMUN(EI)-Anti-SARS-CoV-2 IgA and IgG ELISA test, which contains the S1 subunit of the spike protein (EI S1 IgG or EI S1 IgA), (ii.) the LIAISON® SARS-CoV-2 S1/S2 IgG CLIA test, containing the S1 and S2 domain of the spike protein (DiaSorin S1/S2 IgG), (iii.) the SARS-CoV-2 IgG CMIA from Abbott detecting Anti-nucleocapsid IgG antibodies (Abbott N IgG) and (iv.) the Elecsys® Anti-SARS-CoV-2 ECLIA test from Roche which uses biotinylated and ruthenylated nucleocapsid antigen for the determination of antibodies against SARS-CoV-2 (Roche N Ab).

**Table 1:**
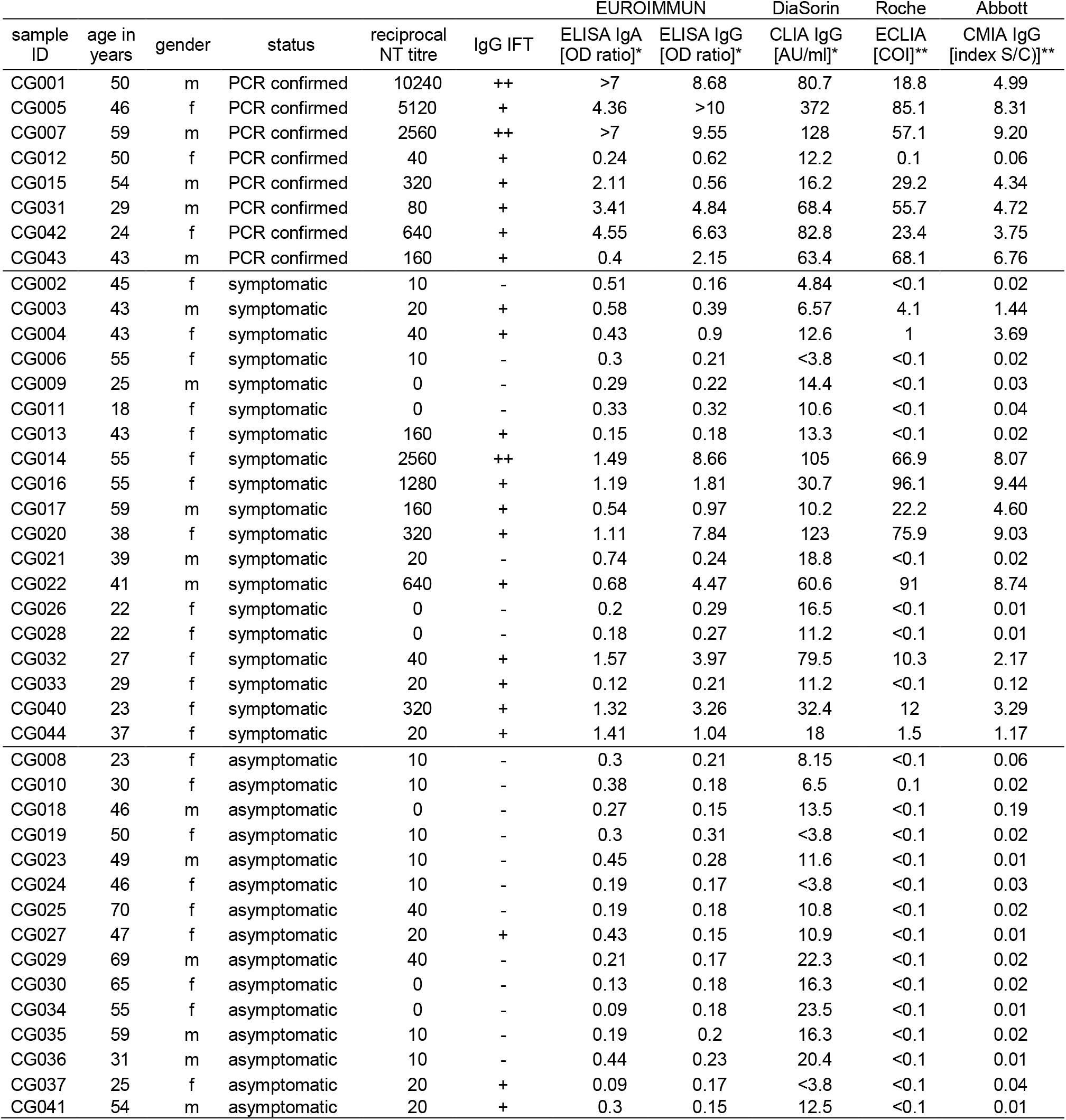
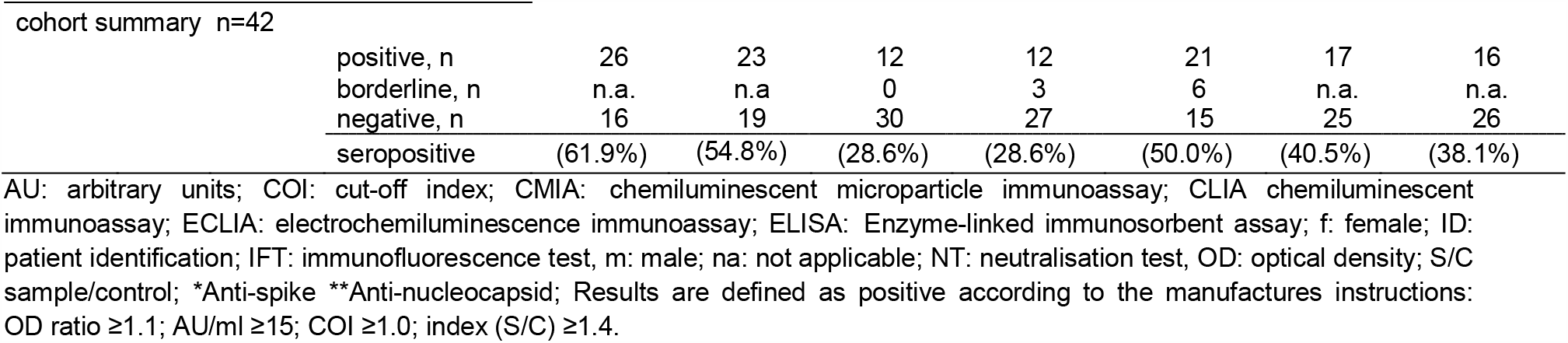
Patient characteristics and serological test results of all Anti-SARS-CoV-2 assays performed in this study, Heinsberg District, Germany, April 2020 (n=42)

Of the 26 SARS-CoV-2 NT-positive sera, 12 were determined positive with the EI S1 IgG or IgA assay, while all 16 NT negative sera have been identified as negative. Of note, 10 of 26 individuals were positive in the EI S1 IgG as well as the EI S1 IgA test (table1). Although the DiaSorin S1/S2 IgG test identified 16 of 26 individuals as positive, 5 of 16 NT negative individuals were tested positive as well. The Abbott N IgG test detected 16 positive individuals in 26 while the Roche N Ab test determined 17 of 26 individuals as positive. In both tests none of the NT negative sera were above the respective cut-off. Thus, the negative agreement between the NT assay and EI S1 IgG or IgA test, the Roche N Ab assay and the Abbott N IgG test were 100%. However, the false-positive rate of the DiaSorin S1/S2 IgG assay was 31.3%. Overall, the EI S1 IgG or IgA test had the lowest sensitivity (46.2% CI 95% [0.355-0.712]; IgA and/or IgG positive 53.8%). The sensitivity of the Abbott N IgG assay as well as the DiaSorin S1/S2 IgG test was 61.54% (CI 95% [0.425-0.776]). Notably, the Roche N Ab assay had the highest sensitivity with 65.4% (CI 95% [0.462 - 0.806]) (table 2). Taken together the N restricted tests showed a better sensitivity compared to S restricted tests. Nevertheless, all currently available test systems missed a large proportion of neutralising sera.

**Table 2:** Performance characteristics of the EUROIMMUN, DiaSorin, Roche and Abbott SARS-CoV-2 antibody platforms, Heinsberg District, Germany, April 2020 (n=26)

|  |  | EUROIMMUN |  |  | DiaSorin | Roche | Abbott |
| --- | --- | --- | --- | --- | --- | --- | --- |
|  |  | S1 IgA | S1 IgG | S1 IgA and/or IgG | S1/S2 IgG | N antibodies | N IgG |
| overall NT positive $\geq 20$ | n/N | 12/26 | 12/26 | 14/26 | 16/26 | 17/26 | 16/26 |
|  | Value | 0.462 | 0.462 | 0.538 | 0.615 | 0.654 | 0.615 |
|  | (95% CI) | 0.288 - 0.645 | 0.288 - 0.645 | 0.355 - 0.712 | 0.425 - 0.776 | 0.462 - 0.806 | 0.425 - 0.776 |

CI: Confidence interval; N: nucleocapsid; NT: neutralisation test; S: spike
For sensitivity calculations of the commercial assays only the NT positive samples ( $\geq 20$ ) were used.

## Correlation of commercial SARS-CoV-2 antibody assay results with neutralisation ability

The neutralisation titre correlated strongly with all spike antigen-based antibody tests (EI S1 IgA r=0.7625; EI S1 IgG r=0.6886; DiaSorin S1/S2 IgG r=0.5641) (figure 2). The weaker correlation of the commercial N-test systems (Abbott N IgG r=0.4579 and Roche N Ab r=0.3523) with the neutralising antibody titres indicates that S-based systems are more likely to be predictive for functional antibodies.

**Figure 2:**
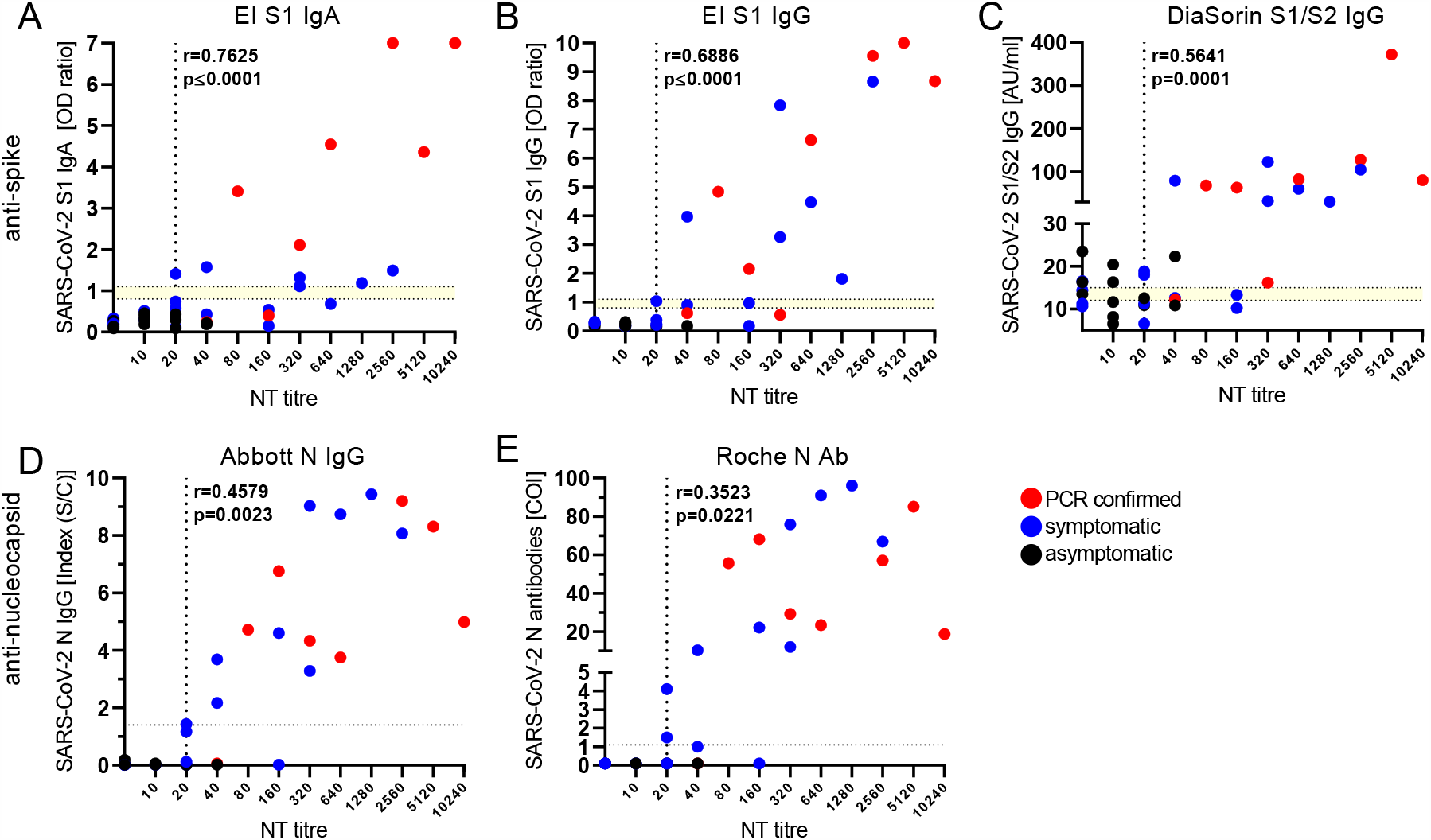
Correlation between commercial SARS-CoV-2 antibody tests and the neutralisation titre, Heinsberg District, Germany, April 2020 (n=42) AU: arbitrary units; COI: cut-off index; EI: EUROIMMUN; N: nucleocapsid; NT: neutralisation test; OD: optical density; r: correlation coefficient; S1: spike domain 1; S2: spike domain 2; S/C: sample/control; SARS-CoV-2: severe acute respiratory syndrome coronavirus 2. The reciprocal of the NT titre is depicted. RT-PCR confirmed SARS-CoV-2 infections are depicted in red and symptomatic individuals in blue. All asymptomatic individuals are displayed in black. (A+B) EUROIMMUN-Anti-SARS-CoV-2 IgA and IgG ELISA (Euroimmun), (C) LIAISON® SARS-CoV-2 S1/S2 IgG (DiaSorin), (D) SARS-CoV-2 IgG CMIA (Abbott), (E) Elecsys® Anti-SARS-CoV-2 ECLIA test (Roche). The dotted lines indicate the cut-off values recommended by the respective manufacturer to determine positive and negative test results. The borderline area if applicable is indicated in yellow and the vertical line represents the positive cut-off of a NT titre ≥20.

## Discussion

In December 2019, a new coronavirus, severe acute respiratory syndrome coronavirus 2 (SARS-CoV-2), emerged in China and its pandemic spread resulted in currently more than 6 million infected people [3-5]. Knowledge of its natural distribution and real infection rates of SARS-CoV-2 in the pre-lockdown settings are urgently needed to predict post-lockdown infection scenarios. To our knowledge, this is the first study comparing the sensitivity of four different currently available commercial antibody tests EUROIMMUN-Anti-SARS-CoV-2 IgA and IgG ELISA, LIAISON® SARS-CoV-2 S1/S2 IgG (DiaSorin) CLIA, the SARS-CoV-2 IgG CMIA from Abbott and the Elecsys® Anti-SARS-CoV-2 ECLIA test from Roche, in a high prevalence setting.

The peculiarity of this study is that the study cohort included 42 individuals who have had contact to the NRW index patient at the end of February 2020, a time of uncontained viral spread since health authorities had not yet taken containment measures. Although only 8 infected individuals were previously tested SARS-CoV-2 PCR positive, we found that 26 of 42 individuals had neutralising antibodies (61.9%) in the NT. This high seroprevalence is consistent with data from a high school in France describing that 40.9% of pupils, teachers and the school staff combined had SARS-CoV-2 antibodies [6]. The study by Streeck et al. [7], sampling 1,007 people from the area where the German Heinsberg outbreak occurred, found an Anti-SARS-CoV-2 seroprevalence in the range of 15%. Importantly, our data suggest that in such a high-prevalence setting a substantial number of convalescent COVID-19 cases may be missed with commercial serological assays. Although we found a high concordance of IFT positive with NT-positive individuals (23 of 26; 88.5% CI95% [0.710-0.960]), the commercially available SARS-CoV-2 antibody assays from four companies evaluated in the current study had a low sensitivity compared to the NT or the IFT. This potentially adds to the difference observed between our study and the one of Streeck et al. as only ELISA IgG seropositive sera were analysed in their NT [7].

Although none of the commercial assays detected more than 65.4% of SARS-CoV-2 NT assay confirmed positive individuals we see a slightly higher sensitivity of nucleocapsid-assays compared to assays using spike which is in line with previous findings [8]. However, since the median time of sera sampling after symptom onset was 43 days, this could not be attributed to an earlier Anti-N response as described by Grzelak et al. [9]. Notably, both N-restricted assays gave no false-positive results in our small cohort even though a higher cross reactivity to human coronaviruses (HCoVs) has been proposed [1]. The sensitivity of the assays as reported by the manufacturers ranged between 93.8% and 100% ≥14 to >21 days post symptom onset. However, critical COVID-19 cases seem to mount a more robust antibody response than non-critical hospitalised patients [10]. Accordingly, all assays detected higher antibody levels in confirmed PCR-positive cases, a group that showed a more severe disease course than the other groups. In turn, the sensitivity might be insufficient for detection of all mild or asymptomatic cases as in this cohort.

In line with previous studies, ELISA and CLIA assays detecting anti-S or anti-N antibodies had a mild to strong correlation with neutralisation titres [11, 12]. The EUROIMMUN-Anti-SARS-CoV-2 IgA and IgG ELISA tests showed the strongest correlation with antibody function (IgA r=0.7625 p≤0.0001; IgG r=0.6886 p≤0.0001) followed by the LIAISON® SARS-CoV-2 S1/S2 IgG assay (r=0.5641 p=0.0001). In the current study, serological assays detecting spike antibodies showed better correlations, which might be due to the fact that the spike protein is the major target for neutralising antibodies for related coronaviruses and proposedly as well for SARS-CoV-2 [13, 14]. Of note, the DiaSorin S1/S2 IgG assay rendered multiple false positive results from NT assay-negative samples and two control samples with known HCoV infection. Therefore, suggesting a cross-reactivity to other endemic HCoVs possibly because the spike S2 subunit is more conserved among HCoVs than the S1 domain [11, 12]. Since neutralising antibody titres in SARS-COV-2 infected individuals varied widely, the EUROIMMUN-Anti-SARS-CoV-2 IgA and IgG assay could be considered for pre-screenings to determine optimal donors for convalescent plasma or estimating the induction of virus-specific neutralising antibodies after vaccination.

Calculation of sensitivity of all commercially available test systems was performed with the NT as reference for past SARS-CoV-2 infection. As 5.7% of hospitalised COVID-19 patients do not generate neutralising antibodies neither at the time of discharge or thereafter [15] we could not exclude the possibility that we potentially missed some SARS-CoV-2 infected individuals. Moreover, since we included volunteers from a high-risk area in this small sample study, the data might not be representative for a low-prevalence setting, which is the current situation in most areas of Europe.

In conclusion, the four commercially available high-throughput assays for the detection of SARS-CoV-2 specific antibodies differed in their sensitivity and their potential to predict the neutralization capacity of patient sera. The N-immunoassays tested here seemed to be more sensitive compared to S1 spike protein assays. However, sensitivity of these assays was still insufficient for detecting all individuals with neutralising capacity. These results should be considered in future population based seroprevalence studies.

## Data Availability

The authors confirm that the data supporting the findings of this study are available within the article and its supplementary materials.

